# Uptake, safety, and effectiveness of inactivated influenza vaccine in patients with inflammatory bowel disease: a nationwide study in the UK using data from the Clinical Practice Research Datalink

**DOI:** 10.1101/2023.09.18.23295713

**Authors:** Georgina Nakafero, Matthew J. Grainge, Tim Card, Christian D. Mallen, Jonathan S. Nguyen Van-Tam, Abhishek Abhishek

## Abstract

**Objective:** We investigated UK wide inactivated influenza vaccine (IIV) uptake in adults with inflammatory bowel disease (IBD), the association between vaccination against influenza and IBD flare, and the effectiveness of IIV in preventing morbidity and mortality.

**Design:** Data for adults with IBD prior to 1^st^ September 2018 were extracted from the Clinical Practice Research Datalink (CPRD) Gold, a database of electronic health records originated during routine care of patients in the UK. It is linked to hospitalization and mortality records. We calculated the proportion of patients vaccinated against seasonal influenza in the 2018-2019 influenza cycle. To investigate vaccine effectiveness, we calculated propensity score (PS) for vaccination and undertook Cox proportional hazard regression with inverse-probability treatment weighting on PS. We employed self-controlled case series (SCCS) to investigate the association between vaccination and IBD flare.

**Results:** Data for 13,631 IBD patients (50.4% male, mean age 52.9 years) were included. Fifty percent were vaccinated during the influenza cycle while 32.1% were vaccinated before influenza virus circulated in the community. Vaccination was associated with a non-significant reduction in hospitalisation for pneumonia (aHR (95%CI) 0.52 (0.20-1.37), including in the influenza active period (aHR (95%CI) 0.48 (0.18-1.27)). Administration of the influenza vaccine was not associated with IBD flare.

**Conclusion:** The uptake of influenza vaccine is low in IBD patients and the majority are not vaccinated before influenza virus circulates in the community. Vaccination with the IIV is not associated with IBD flare. These findings add to the evidence to promote vaccination in patients with IBD.

**Key messages:** *What is already known on the topic:*

- Inactivated influenza vaccine is recommended in people with IBD treated with immune suppressing drugs.
- Concerns about influenza vaccine causing IBD flare and lack of data on the effectiveness of influenza vaccine in people with IBD are barriers to seasonal influenza vaccination in this population.

*What this study adds:*

- The uptake of seasonal influenza vaccination is low in IBD patients.
- Seasonal influenza vaccination is not associated with IBD flare and is more likely to prevent serious complications of influenza in this population.

*How this study might affect research, practice, or policy:*

- This study provides new data on the uptake, effectiveness, and safety of influenza vaccine in people with IBD and adds to the accumulating evidence to promote vaccination in this population.

## Introduction

Inflammatory bowel disease (IBD) is a common immune-mediated inflammatory disease which affects 1.4% of adults in the UK^1^. People with IBD are at an increased risk of influenza and its complications such as hospitalisation and death ^2^. Consequently, annual vaccination with the inactivated influenza vaccine (IIV) is recommended in this population if immunosuppressed due to treatment^3 4^. Despite the recommendation for vaccination, data on the uptake, safety and effectiveness of IIV in IBD are sparse. A cross-sectional survey of 88 IBD patients prescribed immune-suppressing drugs from a UK gastroenterology outpatient clinic found that 61.4% patients were vaccinated against influenza^5^ but in another survey of 89 patients, only 28.1% were vaccinated against influenza during the H1N1 pandemic of 2009^6^. In North America and Europe, influenza vaccine uptake was low, between 28.7% to 34%^7 8^.

In terms of the association between vaccination against influenza and IBD flare, a systematic review reported that 3% patients with IBD experienced an IBD flare after vaccination against influenza – however none of the included studies had a control group and it is unclear whether vaccination against influenza was temporally associated with an IBD flare or whether this was coincidental or ascertainment bias^9^. Similarly, while the IIV was as immunogenic in the IBD population as in healthy adults^10^, the effectiveness of the IIV in people with IBD is has not been evaluated. In immunosuppressed adults with rheumatic disease, IIV protected against influenza-like illness (ILI), hospitalisation due to pneumonia and death due to pneumonia with a vaccine effectiveness (VE) of 30% to 50%^11^. Vaccination was associated with a 9% lower rate of primary-care consultation for joint pain in the 90-days post vaccination^12^.

A lack of knowledge about VE and concerns about safety underlie vaccine hesitancy in people with inflammatory conditions^13^. In order to provide evidence to improve the uptake of vaccination against influenza in IBD, the objectives of this study were to assess the uptake, safety and effectiveness of the IIV in preventing ILI, lower respiratory tract infections (LRTI), pneumonia and death in people with IBD.

## Methods

### Data source

Data from the Clinical Practice Research Datalink (CPRD) Gold were used in this study. Incepted in the year 1987, CPRD Gold is an anonymised longitudinal database of electronic health records of >14 million people in the UK. CPRD participants are representative of the UK population in terms of age, sex, and ethnicity^14^. CPRD includes information on demographics, lifestyle factors, diagnoses stored as Read codes – a coded thesaurus of clinical terms, primary-care prescriptions, and immunisations. Vaccination and date of vaccination are also recorded. The data are enhanced by linkage with hospitalisation (Hospital Episode Statistics (HES)) and mortality records (Office of National Statistics (ONS)).

### Approval

CPRD Research Data governance (Reference 21_000614).

### Study design and period

Cross-sectional, cohort and self-controlled case series study designs were used to examine IIV uptake, safety and effectiveness respectively.

Study period was from 01/09/2018 to 31/08/2019 i.e., start and end of 2018-19 influenza cycle.

### Population

Adults aged _≥_18 years with one or more primary care consultations for IBD and at least one prescription of immune suppressing drugs prior to study start i.e. 1^st^ September 2018 were included. We selected a broad range of drugs that may be used to treat inflammatory bowel disease i.e. 5-aminosalicylates (mesalazine, balsalazide, olsalazine), azathioprine, mercaptopurine, methotrexate, mycophenolate, ciclosporin, tacrolimus, sirolimus for case definition.

### Exposure

Vaccination with the IIV defined using product and Read codes^15^. Dates of vaccination were extracted from CPRD.

### Outcomes

#### Uptake

IIV administration.

#### Effectiveness

[1] primary care consultation for lower respiratory infections (LRTI) defined as primary-care consultation for LRTI and antibiotic prescription occurring on the same date, [2] primary care consultation for ILI defined using Read codes, [3] hospitalisation for pneumonia defined using ICD codes in the linked hospital episode statistics dataset, and [4] all-cause death as described previously^11^. After feasibility assessment, death due to pneumonia was not selected as an outcome due to 11 events.

#### Safety

IBD flare was the outcome of interest. It was defined as a new primary care prescription of corticosteroids or 5-aminosalicylate prescription after a four months gap, a validated approach for definition of flare in IBD^16^. To further improve the positive predictive value, we excluded participants with a record of an alternative indication for corticosteroids on the same date as consultation for IBD flare.

### Covariates

#### Uptake

*A*ge, gender, immune suppressive drug (yes/no) and presence of additional indication for vaccination as specified in the Green book. Briefly, these included chronic heart diseases, chronic respiratory diseases, chronic kidney diseases, chronic liver diseases, chronic neurological diseases, immunosuppression, diabetes and asplenia.

#### Effectiveness

A propensity score (PS) for vaccination was calculated and included as a covariate because participants at risk of influenza are more likely to be vaccinated^17^. The PS included factors that account for confounding by indication, at-risk conditions, Charlson comorbidity index, and health-seeking behaviour as published previously^11^.

#### Safety

As weather might exacerbate IBD^18^, season was a covariate of interest, defined in line with the Meteorological Office: spring (1^st^ March to 31^st^ May), summer (1^st^ June to 31^st^ August), autumn (1^st^ September to 30^th^ November) and winter (1^st^ December to 28^th^ February).

### Follow-up

Participants were followed up from either 1 September 2018 or the date of registration in the current GP-surgery, whichever was the latest to the earliest of date of death, date of last data collection, transfer out of the GP-surgery or 31 August 2019 in the vaccine uptake and safety analyses. In the VE analyses, follow-up was also censored at the outcome date if it occurred earlier than the study end date.

### Statistical analyses

#### IIV uptake

The percentage and 95% CI of participants that received IIV between the start and end of the 2018-2019 influenza cycle, and in-time before influenza circulated in the community (3^rd^ November 2018 as per the weekly Public Health England (PHE), now UK Health Security Agency (UKHSA) bulletins was calculated. The proportion vaccinated was stratified by age (<45, 45-64, _≥_ 65 years), presence of additional indication for vaccination as per the Green book and immune-suppressive drug prescription in the three months immediately prior to the start of influenza cycle. Poisson regression was used to examine the multivariable association between age, sex, immune-suppressing drug use, presence of at-risk condition and receiving the IIV.

#### IIV effectiveness Mean

(standard deviation (SD)), n (%) and standardised difference (*d*) were used to examine covariate balance between vaccinated and unvaccinated participants. PS for vaccination was calculated using logistic regression, treating vaccination status as the dependent variable. Multiple imputation using chained equation was used to impute missing data on smoking, alcohol and body mass index (BMI). Ten imputations were carried out.

Cox regression was used to calculate hazard ratios (HRs) and 95% CIs, combined using Rubin’s rule across the imputed datasets, with vaccination as the exposure of interest. Vaccination was treated as a time varying exposure whereby the time period from date of vaccination was considered as exposed, whilst the period before this contributed to unexposed period. Participants without a vaccination record in the influenza cycle were considered unexposed for the entire duration. Inverse-probability treatment weighting (IPTW) using the PS was performed to account for confounding.

As IIV is most likely to influence outcomes during influenza active periods (IAP), we undertook additional analyses restricted to the IAP. IAPs were defined as per PHE (now UKHSA) reports using information about rates of consultation for ILI, and isolation of the virus from sentinel surveys^19^.

#### IIV safety

Vaccinated participants with _≥_1 IBD flare in the study period were included. The 2018-2019 influenza cycle was divided into baseline, pre-vaccination and post-vaccination periods. The baseline extended from 1^st^ September 2018 to 15 days pre-vaccination, and from 90 days post-vaccination to earliest of 31^st^ August 2019, date of leaving GP surgery, date of death, or latest date of data collection. The exposed period extended from vaccination to 90 days later and, was further categorised as 0-14 days, 15-30 days, 31-60 days, and 61-90 days post-vaccination. The first categorization of exposed period was at 14-days post-vaccination as it takes approximately 2 weeks for the serological response and, this period of immune reconstitution would be most strongly associated with disease activity if such an association were to exist. The 14-day period immediately preceding vaccination was excluded from the baseline period to minimise confounding due to healthy vaccinee effect which results in people seeking vaccination only when they are well. A Poisson model conditioned on the number of events adjusted for the seasons as categories defined in line with the Meteorological Office description was fitted to calculate incidence rate ratios (aIRR) and 95% CI for each exposure period compared to the baseline period. Data management and analysis were performed in Stata v17, Stata Corp LLC, Texas, USA.

### Patient and public involvement

Patient and members of the public were involved in selecting the research question. They advised us to use readily available datasets instead of undertaking an expensive primary study.

### Results

Data were available for 13,631 IBD patients (Figure 1). Of these, 50.4% were male, mean (SD) age was 52.9 (17.4) years, 45.2% were current or previous smokers, 34.2% had at least one additional at-risk condition for vaccination, and 50.1% and 38.4% were previously vaccinated against influenza or pneumococcal vaccinations respectively (Table 1).

**Figure 1:**
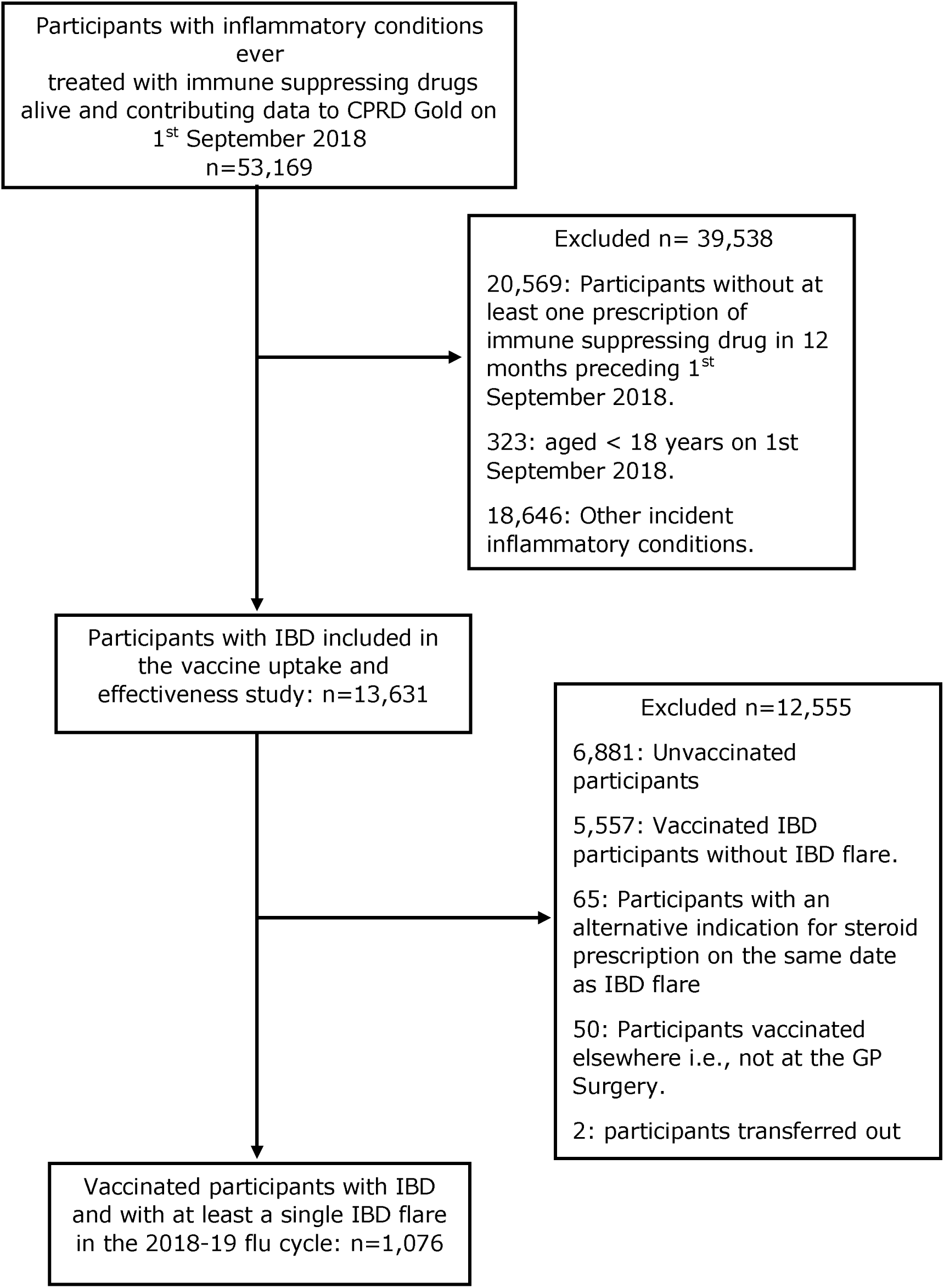
Population selection criteria for IIV uptake. CPRD: Clinical Practice Research Datalink.

**Tables 1:** Cohort characteristics (n=13,631).

| Variable | Distribution |  |
| --- | --- | --- |
| Continuous variables; | mean (SD) |  |
| Age | 52.9 (17.4) | *Number<br>(percent<br>). |
| Body mass index (BMI) | 27.1 (5.8) |  |
| Missing BMI values | 1,553 (11.4) * |  |
| Charlson's comorbidity index | 0.9 (1.4) |  |
| Index of Multiple deprivation | 3.1 (1.41) |  |
| Number of prescriptions<sup>‡</sup> | 45.8 (57) |  |
| Number of consultations<sup>‡</sup> | 12.1 (10.1) |  |
| Number of hospitalisations<sup>‡</sup> | 0.1 (0.5) |  |
| Categorical variables; | n (%) |  |
| Male sex | 6869 (50.4) |  |
| *Smoking status* |  |  |
| Non-smoker | 7225 (53) |  |
| Current smoker | 1546 (11.3) |  |
| Ex-smoker | 4623 (33.9) |  |
| Missing | 237 (1.7) |  |
| *Alcohol consumption (units/week):* |  |  |
| Non-drinker | 1972 (14.5) |  |
| Low drinker (1-14) | 7173 (52.6) |  |
| Moderate (15-21) | 723 (5.3) |  |
| Hazardous (> 21) | 757 (5.6) |  |
| Former drinker | 803 (5.9) |  |
| Missing | 2203 (16.2) |  |
| Nursing Home resident | 86 (0.6) |  |
| Previous influenza vaccination | 6835 (50.1) |  |
| Previous pneumococcal vaccination | 5228 (38.4) |  |
| Diabetes | 1232 (9.0) |  |
| Immunosuppression | 200 (1.5) |  |
| Chronic kidney disease | 927 (6.8) |  |
| Chronic respiratory disease | 2746 (20.2) |  |
| Chronic heart disease | 586 (4.3) |  |

#### IIV uptake

Vaccine uptake during the entire 2018-2019 influenza cycle and in the time of influenza virus circulation in the community was 49.52% and 32.11% respectively. Vaccine uptake (95% CI) was 32.56% (31.50-33.64%) in the low-risk groups defined as the under 65s and patients without additional at-risk conditions, and 69.45% (95% CI 68.30-70.58%) in the high-risk group defined as patients aged _≥_ 65 years or those with additional at-risk condition. Increasing age, female sex, at-risk condition, and immunosuppression use were independently associated with IIV uptake (Table 2).

**Table 2:** Percentage and risk factors of inactivated influenza vaccine uptake in patients with inflammatory bowel disease during the 2018-19 flu cycle.

|  |  | Vaccinated in the entire flu cycle |  | Vaccinated in-time before flu circulation |  | Incidence rate ratios (IRR)<sup>‡</sup> |  |
| --- | --- | --- | --- | --- | --- | --- | --- |
|  |  | n | Percent (95% CI) | n | Percent (95% CI) | Crude IRR (95% CI) | Adjusted IRR (95% CI) |
| Overall |  | 6,750 | 49.52 (48.68,50.36) | 4,377 | 32.11 (31.33,32.90) | - | - |
| Age, years |  |  |  |  |  |  |  |
|  | < 45 | 1,613 | 33.47 (32.15,34.82) | 994 | 20.63 (19.51,21.79) | 1 | 1 |
|  | 45-64 | 2,224 | 43.22 (41.87,44.58) | 1,461 | 28.39 (27.18,29.64) | 1.29 (1.21,1.38) | 1.34 (1.25,1.43) |
|  | ≥65 | 2,913 | 79.46 (78.12,80.74) | 1,922 | 52.43 (50.81,54.04) | 2.37 (2.23,2.52) | 2.38 (2.23,2.54) |
| Sex |  |  |  |  |  |  |  |
|  | Male | 3,263 | 47.50 (46.32,48.69) | 2,097 | 30.53 (29.45,31.63) | 1 | 1 |
|  | Female | 3,487 | 51.57 (50.38,52.76) | 2,280 | 33.72 (32.60,34.85) | 1.09 (1.03,1.14) | 1.10 (1.05,1.15) |
| Additional at-risk conditions* |  |  |  |  |  |  |  |
|  | Absent | 3,599 | 40.14 (39.13,41.15) | 2,319 | 25.86 (24.97,26.78) | 1 | 1 |
|  | Present | 3,151 | 67.56 (66.20,68.89) | 2,058 | 44.13 (42.71,45.55) | 1.68 (1.60,1.77) | 1.38 (1.31,1.45) |
| Immune-suppressing drugs<sup>†</sup> |  |  |  |  |  |  |  |
|  | 5-aminosalicylates | 4,413 | 45.30 (44.32,46.29) | 2,877 | 29.53 (28.64,30.45) | 1 | 1 |
|  | Immunosuppressants<sup>‡</sup> | 2,337 | 60.08 (58.53,61.61) | 1,500 | 38.56 (37.04,40.10) | 1.33 (1.26,1.39) | 1.57 (1.50,1.66) |

<sup>‡</sup> Incidence rate ratios for vaccine uptake in the entire flu cycle; <sup>†</sup> latest prescription with 12 months prior to start of 2018-19 flu cycle (01/09/2018); <sup>‡</sup> Methotrexate, leflunomide, thiopurines, ciclosporin, mycophenolate, tacrolimus, and sirolimus. \*There were no patients in with asplenia.

#### IIV effectiveness

Primary care consultation for LRTI requiring antibiotics, ILI, hospitalisation due to pneumonia and all-cause death occurred in 294, 38, 45 and 465 participants at an incidence rate (95%CI) of 23.35 (20.83,26.18), 2.98 (2.17,4.10), 17.47(13.05,23.40), 36.46 (33.29,39.92) per 1,000 person-years respectively. PS was calculated after imputation of 1,553 (11.4%), 237 (1.7%) and 2,203 (16.2%) missing values on smoking, alcohol consumption and BMI respectively. Covariate balance between the influenza unvaccinated and vaccinated IBD patients was achieved following IPTW on PS (Table 3).

**Table 3:** Covariate balance before and after inverse probability of treatment weighting (IPTW) using the propensity score. *.

| | Vaccinated<br>(n=6,750) | Unvaccinated<br>(n=6,881) | $d^{\dagger}$ before<br>IPTW | $d^{\dagger}$ after<br>IPTW |
| --- | --- | --- | --- | --- |
| Continuous covariates; mean (SD) |  |  |  |  |
| Age | 59 (18) | 47 (15) | 0.722 | -0.067 |
| Body mass index | 27.4 (6) | 26.5 (5.7) | 0.166 | -0.025 |
| Charlson's comorbidity index | 1.22 (1.61) | 0.49 (1) | 0.552 | -0.055 |
| Index of Multiple deprivation | 3.1 (1.4) | 3.1 (1.4) | 0.027 | 0.004 |
| Number of prescriptions<sup>‡</sup> | 61.5 (66.8) | 30.4 (39.7) | 0.565 | -0.087 |
| Number of consultations<sup>‡</sup> | 14.8 (11) | 9.4 (8.3) | 0.546 | -0.035 |
| Number of hospitalisations<sup>‡</sup> | 0.1 (0.5) | 0.1 (0.4) | 0.076 | -0.050 |
| Categorical covariates; n (%) |  |  |  |  |
| Male | 3263 (48.3) | 3606 (52.4) | 0.081 | 0.002 |
| *Smoking status* |  |  |  |  |
| Current smoker | 639 (9.5) | 937 (13.6) | 0.130 | 0.008 |
| Ex-smoker | 2652 (39.3) | 2031 (29.5) | 0.207 | -0.004 |
| *Alcohol consumption (units/week):* |  |  |  |  |
| Low drinker (1-14) | 4190 (62.1) | 4382 (63.7) | 0.033 | 0.009 |
| Moderate (15-21) | 408 (6) | 462 (6.7) | 0.027 | 0.005 |
| Hazardous (> 21) | 380 (5.6) | 516 (7.5) | 0.076 | -0.003 |
| Former drinker | 554 (8.2) | 375 (5.5) | 0.109 | -0.012 |
| Nursing Home | 59 (0.9) | 27 (0.4) | 0.061 | -0.024 |
| Previous influenza vaccination | 5648 (83.7) | 1187 (17.3) | 1.777 | -0.011 |
| Previous pneumococcal vaccination | 3988 (59.1) | 1240 (18) | 0.930 | -0.016 |
| Diabetes | 960 (14.2) | 272 (4) | 0.363 | -0.030 |
| Immunosuppression | 150 (2.2) | 50 (0.7) | 0.124 | -0.032 |
| Chronic kidney disease | 735 (10.9) | 192 (2.8) | 0.325 | -0.058 |
| Chronic respiratory disease | 1677 (24.8) | 1069 (15.5) | 0.233 | 0.002 |
| Chronic heart disease | 489 (7.2) | 97 (1.4) | 0.290 | -0.001 |

<sup>†</sup>Standardised difference; <sup>‡</sup> In previous 12 months, \*data from one imputed dataset.

The IIV protected from all-cause death (aHR (95% CI): 0.73 (0.55,0.97). Similarly, IIV reduced the risk for hospitalisation for pneumonia, however, the confidence interval was wide due to a low number of events (aHR (95%CI) 0.52 (0.20,1.37). There was no statistically significant association between IIV and primary care consultation for LRTI (aHR(95%CI): 1.42 (0.98 to 2.06)) and ILI (aHR (95%CI): 1.42 (0.59 to 3.41)). These findings remained unchanged when restricted to the IAP (Table 4).

**Table 4:**
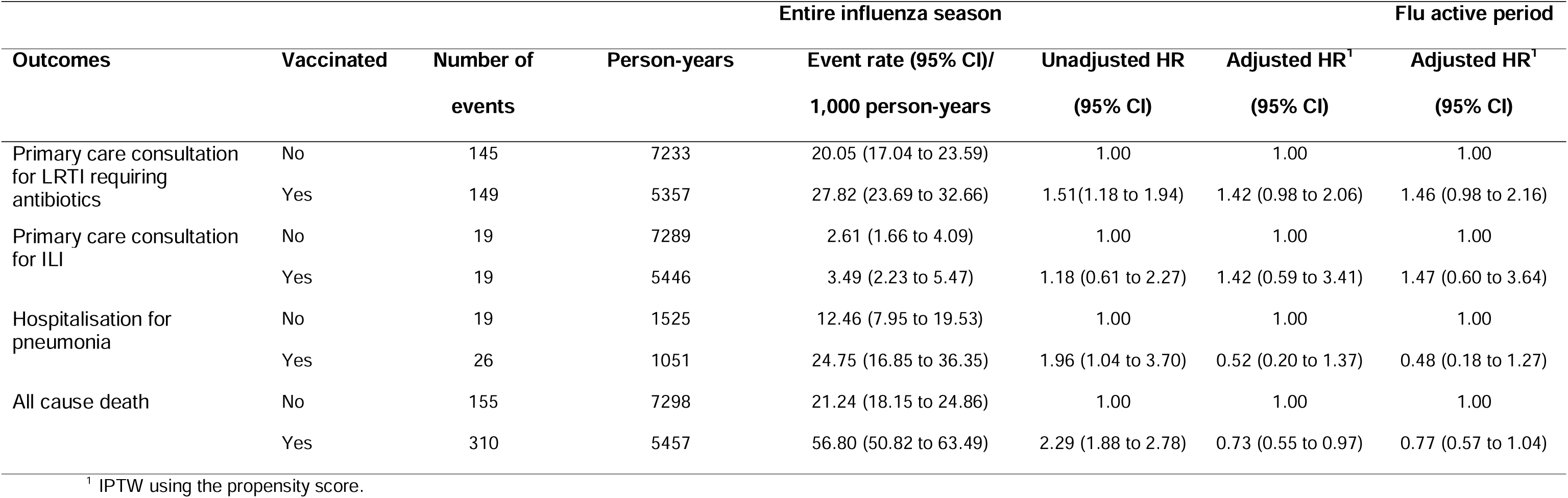
Inactivated influenza vaccine effectiveness in people with inflammatory bowel disease.

#### IIV safety

Data for 1,076 vaccinated participants with IBD flare were included in the analysis (Figure 1). The majority of participants were female (53.5%) and their mean (SD) age was 55 (17) years. 581 (54%) had UC, 339 (31.5%) had Crohn’s disease, 156 (14.5%) had IBD without any specific coding for subtype. 906 (84.2%), 162 (15.1%) and 8 (0.7%) participants had one, two, and three IBD flares respectively. 19 participants (1.8%) did not contribute data for the entire follow-up period due to death (n = 7 (0.7%)) or transfer out of GP practice (n = 12 (1.1%)). Vaccination against flu was not associated with IBD flares in the 90-days post vaccination (Table 5).

**Table 5:** The association between Flu vaccination and IBD flare.

| Risk period (days) | Events (n) | IRR (95%CI) | Adjusted IRR (95%CI) * | p-value |
| --- | --- | --- | --- | --- |
| Baseline | 884 | 1.00 | 1.00 |  |
| 15 days pre-vaccination | 61 | 1.17 (0.90,1.51) | 1.13 (0.84,1.52) | 0.431 |
| Post vaccination intervals |  |  |  |  |
| 0 - 90 days | 309 | 0.99 (0.87,1.12) | 0.68 (0.46,1.02) | 0.060 |
| 0 - 14 days | 51 | 0.97 (0.73,1.29) | 0.92 (0.72,1.36) | 0.941 |
| 15 - 30 days | 55 | 1.05 (0.80,1.38) | 0.99 (0.72,1.36) | 0.941 |
| 31 - 60 days | 99 | 0.95 (0.77,1.17) | 0.87 (0.62,1.21) | 0.409 |
| 61 - 90 days | 104 | 1.00 (0.81,1.22) | 0.90 (0.63,1.29) | 0.576 |

\* adjusted for Season.

## Discussion

This large nationwide wide study found that the IIV uptake is low in people with IBD, even among those with additional indication for vaccination, and a substantial proportion of vaccinated patients did not receive optimally timed vaccination before the influenza virus began to circulate in the community. However, the vaccine did not produce an excess of IBD flares and was likely to protect against severe outcomes of influenza specifically hospitalization for pneumonia and all cause death. The latter could be due to unmeasured confounding and is not likely to be a causal effect.

Our estimate of 49.5% IIV uptake is comparable to those observed in previous questionnaire surveys. In North America and Europe, influenza vaccine uptake ranged between 28.1% and 61.4% ^7 856^. Unlike prior studies limited by the small sample size and use of survey questionnaire prone to recall bias, our findings are from CPRD, a large primary care database, suitable for this study as vaccination in the UK is undertaken in primary care. As reported in UK autoimmune rheumatic diseases, the vaccine uptake was higher in women than in men and increased with age^15 20^. Unsurprisingly, presence of comorbidities and immunosuppressive drug treatment significantly associated with vaccine uptake as reported in rheumatic diseases^20^.

Vaccination with the IIV was not associated with an increased risk of IBD flare in this study. This is consistent with uncontrolled data from previous studies^9 21 22^. In an observational study of 575 IBD patients on immunomodulators or anti-TNFs vaccinated between November 2009 and March 2010 in 14 European countries, H1N1 vaccine was found to be well tolerated in terms of disease control with no flares in over 96% patients^22^. In a randomised controlled trial of 137 subjects with IBD on maintenance infliximab therapy allocated to receive the 2012/2013 IIV at different time-points with respect to the infliximab infusion, adverse effects lasting longer than 24 hours were infrequent and there were no severe adverse effects requiring medical attention^21^. Similarly, COVID-19 vaccine did not associate with IBD flares^23^.

In this study, IIV showed a tendency to protect against pneumonia. It is difficult to compare these results with those of previous studies, since to our knowledge, this is the first study to assess clinical effectiveness of seasonal IIV in IBD. Immunologic studies in children and adults with IBD found IIV to generally induce appropriate immune response to influenza ^24 25^. However, when patients are receiving immunosuppressive therapies with combined thiopurines and anti-TNF-alpha agents, serologic response to vaccines is lower compared to monotherapy or non-immune suppressing treatment^9 25 26^. Nonetheless, even a blunted vaccination response is thought to be of benefit ^27^ and immunological correlates of protection against influenza remain poorly understood^28^. Although the 48% effectiveness in reducing hospitalisation for pneumonia lacked statistical significance in this IBD population, in our previous study of rheumatic disease patients, IIV associated with reduced ILI and complications of influenza including hospitalisation for pneumonia and all-cause death at 48% and 59% VE respectively^11^. Another study reported 35% VE for hospitalisation due to septicaemia, bacteraemia or viremia and 38% VE for all-cause mortality with IIV in people with Rheumatoid Arthritis^29^. Estimates for the associations between IIV and primary care consultation for LRTI and ILI and results from restricted analyses to influenza active periods, lacked statistical significance in this IBD population unlike in our previous study of the effectiveness of IIV in a similar immunocompromised population^11^. The lack of statistical significance may be attributed to the low number of outcomes in current study. Additionally, PHE (now UKHSA) VE data for 2018-19 also showed poor protection against infection in adults for influenza A(H3N2)^28^. Primary care consultation for LRTI was selected as an outcome because bacterial chest infection is a complication of influenza. However, there was no evidence of a protective effect on this outcome which could be due to. risk-averse antibiotic prescription to potentially immunosuppressed people in primary care.

Strengths of this study include the generalisability of its findings to patients with IBD in the UK due to its data source, CPRD which is representative of over 98% of the UK population registered with a GP surgery^31^. We used a combination of diagnostic and prescription codes to identify people with IBD, increasing the validity of our case definition. Studies of VE are biased due to confounding by indication and healthy user bias, but we attempted to account for this using PS for vaccination and employing inverse probability treatment weighting on the PS in the Cox regression analysis. Nevertheless, it is possible that our results are influenced by unmeasured confounding. Our use of a self-controlled case series methodology that is widely used in vaccine safety studies^32^, ensures that non time-dependent between-person confounding was excluded because participants were compared only with themselves at different time points^33^. There was no selection bias because all patients who had received both vaccination and experienced an IBD flare were included in the IIV safety analysis.

However, this study has several limitations. First, we could not include laboratory confirmed influenza as an outcome as routine viral testing is not conducted for patients presenting to GPs. For this reason, we included ILI as an outcome – this could include many other viral illnesses that are not preventable by vaccination against influenza. Second, we were unable to assess the association between vaccination and death due to pneumonia because of only 11 events. Third, vaccinations occurring outside of the GP surgery are not recorded in the CPRD. This biases the VE results towards null rather than inflating estimates and could explain the lack of significant protective effect on respiratory morbidity and mortality. Fourth, we were unable to assess the impact of biologics on IIV safety because these agents are not recorded in the CPRD. We see no reason though to expect more extreme immunologically driven side effects in these groups given IIV has been shown to be less immunogenic with biologic use^21^. Fifth, because our definition of IBD flare was based on corticosteroid or 5-ASA prescription, minor flares not needing drug treatment were excluded. It is possible that there may be an association with minor flares that were not ascertained; on balance, such effects would be unlikely to greatly discourage vaccination uptake and it is the more significant flares that we have studied which are of primary concern. Finally, because we considered a single influenza season, our study is limited by low power.

In conclusion, this study provides the first UK-wide population-based evidence that the uptake of vaccination against influenza is low in people with IBD and that vaccination does not occur in time before the virus circulates in the community. Vaccination against influenza does not associate with IBD flare. Our VE findings together with results from immunological studies, and other immunocompromised populations suggested that the IIV offers protection from influenza and its complications without an associated risk of IBD flare. These data should be used to promote vaccination in IBD population.

## Data Availability

Due to CPRD licencing rules, we are unable to share data used in this study with third parties. The data used in this study may be obtained directly from the CPRD.

https://cprd.com/

## Author contribution

The study was conceived by Prof Abhishek. All authors were involved in the design of the study. The analysis was carried out by Dr Nakafero. All authors edited the first and all subsequent drafts and approved the final draft for submission.

## Data sharing statement

Data used in the study are from the Clinical Practice Research Datalink (CPRD). Due to CPRD licencing rules, we are unable to share data used in this study with third parties. The data used in this study may be obtained directly from the CPRD. Study protocol is available from www.cprd.com.

